# Shared Genetic Susceptibility between Abdominal Aortic Aneurysm and Cardiometabolic Traits

**DOI:** 10.1101/2023.12.05.23299523

**Authors:** Shufen Zheng, Philip S. Tsao, Cuiping Pan

## Abstract

Abdominal aortic aneurysm (AAA) presents abnormal metabolism and co-occurs with cardiometabolic disorders, suggesting a shared genetic susceptibility. We investigated this commonality leveraging recent GWAS studies of AAA and 32 cardiometabolic traits (CMTs). Significant genetic correlations are found between AAA and 21 CMTs, among which are causal relationship with coronary artery disease, hypertension, lipid traits, and blood pressure. For each trait pair, we identified shared causal variants, genes, and pathways, which revealed cholesterol metabolism and immune responses were the shared most prominently. Additionally, we uncovered the tissue and cell type specificity in the shared signals, with strong enrichment across traits in liver, arteries, adipose tissues, macrophages, adipocytes, and fibroblasts. Finally, we leveraged drug-gene databases and identified several lipid-lowering drugs and antioxidants with high potential to treat AAA with comorbidities. Our study provides insight into the shared genetic mechanism for AAA and cardiometabolic traits and potential targets for pharmacological intervention.

## Introduction

Abdominal aortic aneurysm (AAA), defined as focal dilation of the abdominal aorta by 50% or reaching ≥ 30 mm in diameter, is a complex vascular disease affecting 3-9% population aged over 65 years ^1, 2^. It is asymptomatic in early disease stages, with most AAA discovered by incidental imaging or screening protocols. Once reaching 55 mm, the risk of rupture increases to 10% ^3^. Among ruptured patients, a mortality rate as high as 80% was observed ^4^, rendering AAA a leading cause of death.

Pathologically, AAA is characterized by remodeling and degradation of extracellular matrix, apoptosis of smooth muscle cells, luminal thrombosis, and chronic inflammation ^1^. Plaques consisting of lipids, blood cells and other plasma substances accumulate around the lesion sites, with abundant infiltration of innate and adaptive immune cells both in the thrombus and the arterial wall ^2^. Meanwhile, metabolic homeostasis is perturbed, resulting in enhanced glycolysis in the aortic wall^5^ and altered serum levels of amino acids and lipids ^6, 7, 8^. For example, circulating total cholesterol, low-density lipoprotein cholesterol (LDL-C), triglycerides, and sulfur amino acids are elevated, whereas high-density lipoprotein cholesterol (HDL-C) and phosphatidylcholines are reduced. These changes resemble numerous other cardiovascular diseases (CVDs), such as coronary artery disease (CAD), myocardial infarction (MI), and peripheral arterial disease ^9^. Indeed, atherosclerosis occurs in 25-55% AAA patients ^10^, and known risk factors of AAA including male sex, age, smoking, hypercholesterolemia, hyperlipidemia, and hypertension ^11^, are widely shared among CVDs.

AAA is highly heritable, with an estimated 70% heritability by family and twin studies ^12, 13^. In fact, high heritability is generally observed in cardiometabolic disorders ^14, 15^, rendering genetic studies a valuable tool to decipher the disease mechanisms ^16^. Genome-wide association studies (GWAS), particularly those performed in recent years with large sample sizes, have uncovered single nucleotide variants (SNVs) associated with many complex diseases ^17^. A recent meta-GWAS of AAA examined 39,221 cases and 1,086,107 controls, resulting in 141 susceptible loci ^18^, a several-fold increase in disease loci compared to earlier studies ^19, 20^. Similarly, recent GWAS provided comprehensive mutation profiles for dozens of cardiometabolic traits (CMTs), bringing the disease understanding to a new level.

In this study, we leverage these large GWAS data to study how AAA relates to other CMTs. We aim to construct a map of comprehensive relationship, as well as to provide details such as shared SNVs, genes, and pathways in the cell type and tissue context. Importantly, this comorbidity landscape offers valuable information for prioritizing drugs that target shared genes.

## Results

### GWAS datasets

We obtained GWAS summary statistical data for 18 cardiometabolic diseases (CMDs) including AAA, 15 metabolic traits, and 6 immune cell traits (Fig. 1A). These traits distributed over a broad spectrum of cardiac and metabolic functions, including heart functions, vascular circulation, glucose metabolism, lipid metabolism, and immunity. Most of the CMDs were studied in more than 10,000 case samples, whereas metabolic traits and immune cells were measured in a minimum of 560,000 individuals. Although European ancestry was dominant, many studies included various ancestral groups. Furthermore, the number of interrogated genotypes ranged between 4.5-52 million, and the significant SNVs (*P* < 5 × 10^-8^) were ample (Supplementary Table 1). Overall, these datasets present a state-of-the-art discovery power for common SNVs-based genetic susceptibility to cardiometabolic disorders. Around these datasets, we designed analysis modules to elucidate the shared genetic architecture of AAA and CMTs, including shared SNVs, genes, pathways, tissues, and cell types (Fig. 1B). Coherent signals from various analyses are found and presented below.

**Fig. 1.**
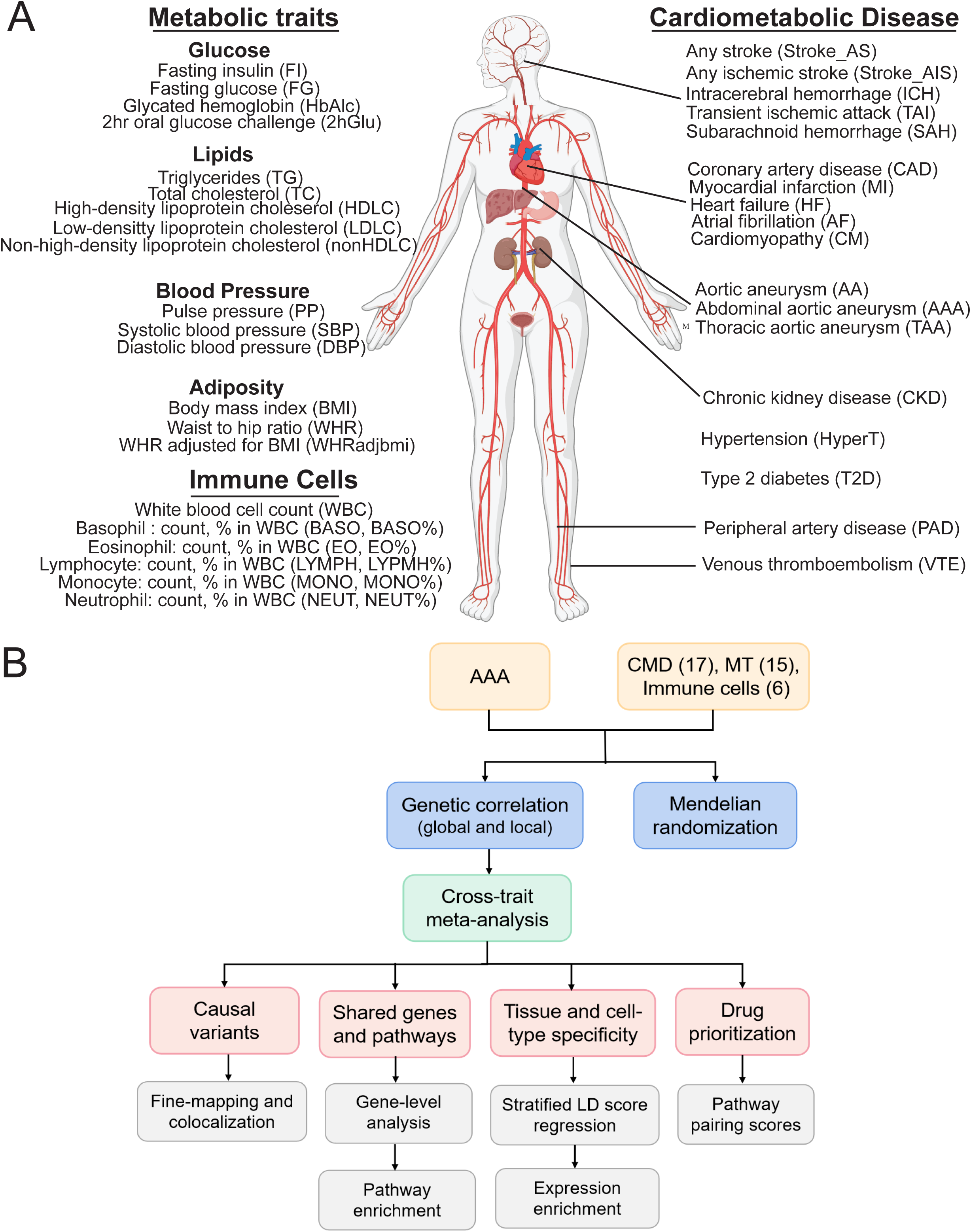
Interrogation of genetic components between abdominal aortic aneurysm (AAA) and the traits related to cardiometabolism (CMTs). **A** Traits and diseases in this study include 18 cardiometabolic diseases, 15 metabolic traits, and 6 immune cell traits. This graph was created via https://www.biorender.com/. **B** Analysis modules included computing genome-wide and local genetic correlations, inferring causality between AAA and the traits by bidirectional Mendelian randomization, identifying shared causal loci, genes, and pathways, discovering tissues and cell being impacted the most by the shared signals, and prioritizing drugs for treating AAA comorbidities. CMD: cardiometabolic diseases, MT: metabolic traits.

### Genetic correlation

Genome-wide correlations computed by LDSC ^21^ suggest positive correlations between AAA and 20 CMTs (Fig. 2A). The highest correlated traits are aortic aneurysms, followed by numerous diseases including MI, CAD, peripheral artery disease, subarachnoid hemorrhage, and heart failure (r_g_ >= 0.3, *P* < 1 × 10^-10^). Compared to the disorders, the physiological traits display weaker correlations, with lipids, adiposity, blood pressure, and glucose traits in the descending order. Only HDL-C presented negative correlation with AAA (r_g_ = -0.25, *P* = 7.61 × 10^-32^). Immune cell counts and percentages did not correlate with AAA, thus were excluded from subsequent analyses.

**Fig. 2.**
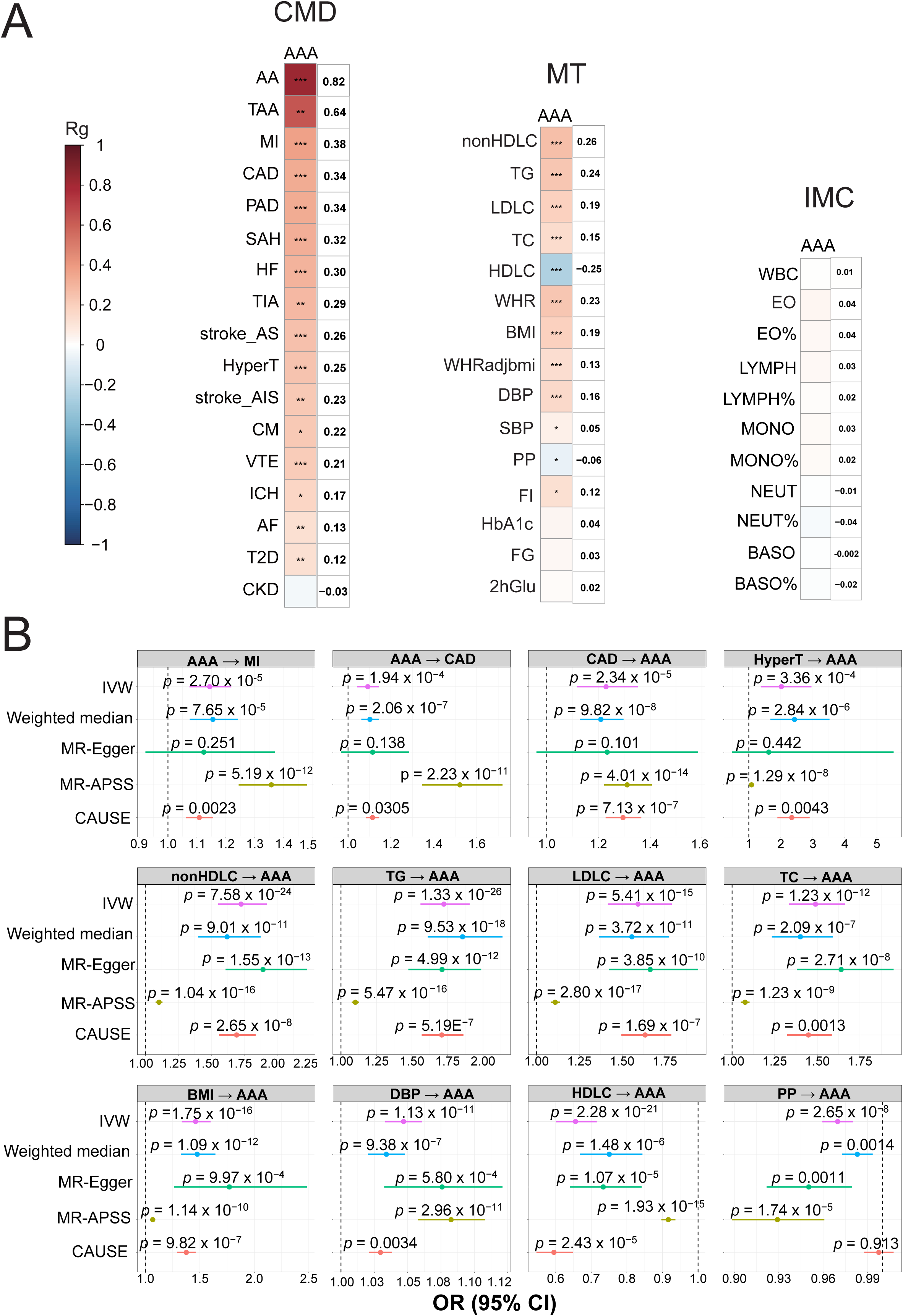
Genetic correlation and causal inference between AAA and CMTs. **A** The heatmap represents the genetic correlation r_g_ calculated in LDSC, with the color scale indicates the strength of the correlation, and the r_g_ value displayed next to the heatmap. The * marks the statistical significance. *: *P* < 0.0016 (Bonferroni-corrected *P* value threshold); **: *P* < 5 × 10^-8^ (genome-wide *P* value threshold). **B** Causal inference by two-sample Mendelian Randomization with five methods. The color bars show causal pairs with the odds ratios +/- 95% confidence intervals and *P* values are depicted above the bars. CMD: cardiometabolic diseases, MT: metabolic traits. IMC: immune cell traits.

We also computed genetic correlation by functional elements. Repressors, enhancers and promoters tend to have stronger than the genome-wide correlations (Supplementary Fig. 1), suggesting transcription regulation is genetically shared.

### Causal Inference

Many cardiometabolic disorders share risk factors, rendering genetic correlation a result of complex pleiotropic effects. Mendelian Randomization (MR) overcomes the confounding factor issue and provides causal inference. We conducted bidirectional MR using several models and found a mutual causality between AAA and CAD (Fig. 2B) . Furthermore, AAA was suggested causal to MI. Reversely, 10 traits were inferred as causal to AAA, among which hypertension had the greatest causal effect (OR = 2.01, *P* = 3.36 × 10^-4^), followed by lipid and adiposity traits (OR = 1.46-1.73, *P* < 1.24 × 10^-12^), and CAD (OR = 1.23, *P* =2 .34 × 10^-5^). Diastolic blood pressure displayed a weak causality (OR = 1.05, *P* = 1.13 × 10^-11^). Conversely, HDL-C (OR = 0.65, *P* = 2.28 × 10^-21^) and pulse pressure (OR = 0.97, *P* = 2.65 × 10^-8^) were causally protective against AAA. Note that no apparent horizontal pleiotropy was detected as the intercept of MR-Egger did not significantly deviate from zero (Supplementary Table 2).

### Cross-trait loci and causal variants

By cross-trait meta-analysis by MTAG (Multi-Trait Analysis of GWAS) ^22^ and CPASSOC (Cross-Phenotype Association Analysis) ^23^, we identified 203 SNVs collectively shared by the 21 trait pairs (Supplementary Table 3). Overall, AAA shares the largest number of SNVs with CAD (N = 46), followed by lipid traits (about 20 - 40 SNVs) (Supplementary Fig. 2).

To derive causal SNVs, we applied FM-summary ^24^ for fine-mapping and derived a 99% credible set (Supplementary Table 4). We then colocalized these SNVs across traits by Coloc ^25^ and derived a total of 177 causal variants shared by two traits (Supplementary Table 5). We also applied HyPrColoc ^26^ and derived 47 causal variants shared by multiple traits (Fig. 3A). Among the 47 shared causal variants, only four were local lead SNVs (Fig. 3B), i.e., having the smallest GWAS *P* values, whereas the rest were located near the lead SNVs, suggesting the importance of fine-mapping.

**Fig. 3.**
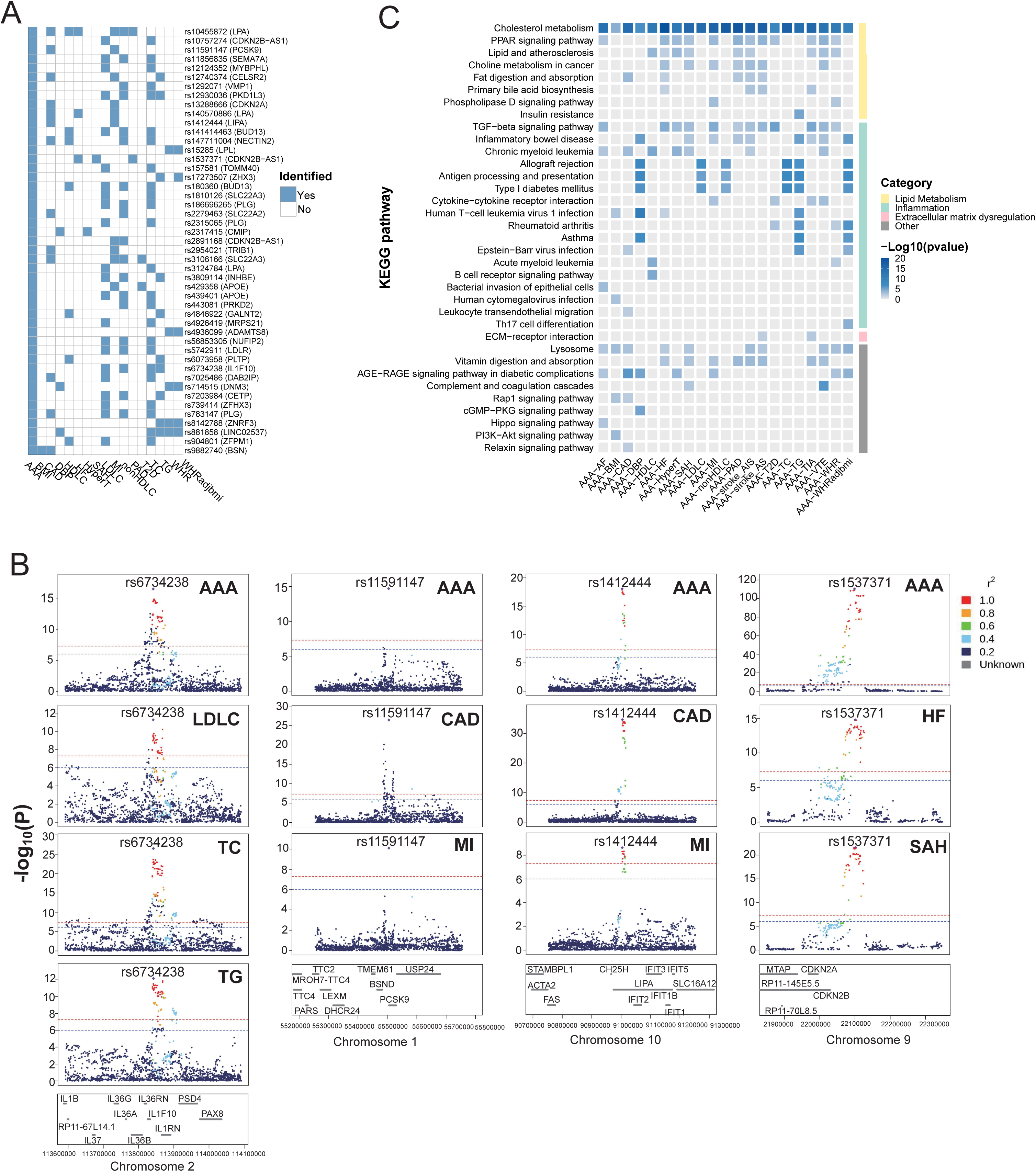
The overall landscape of the pleiotropic associations across AAA and CMTs. **A** 47 causal variants are shared by multiple traits, as identified by HyPrColoc. **B** LocusZoom plots of four causal variants for AAA and multiple other CMTs. These variants are also the lead SNPs in in the interrogated regions. **C** KEGG pathway enrichment of the shared genes between AAA and CMTs, with representation organized by biological mechanisms. Only the top 15 enrichened pathways passing *P* < 0.05 in each cross-trait pair were included.

We observed clusters of shared SNVs, both causal and non-casual, on lipid-related genes (Supplementary Table 5). For example, *LPA* was annotated to 9 shared SNVs, of which 3 were causal to multiple trait, including rs10455872 ^27^ in the intronic region which was causal to 6 AAA trait pairs, and rs140570886 ^28^ and rs3124784 which were shared among 4 and 3 trait pairs, respectively. Similarly, *CDKN2B-AS1* was annotated to 8 shared SNVs, including rs10757274 ^29^, therein 3 were causal and collectively shared among 7 trait pairs,. We also rediscovered rs12740374 ^30^ on *CELSR2* and rs11591147 ^31^ on *PCSK9.* Lastly, several shared causal SNVs were proximal to lipid-related genes, including *CETP*, *BUD13*, *TRIB1*, *LPL*, and *APOE*, all of which encode lipid regulators and have been associated with CMDs ^32, 33, 34, 35, 36^.

### Shared genes and pathways

We adopted four approaches, TWAS ^37^, SMR ^38^, MAGMA^39^, and GCTA ^40^ to infer shared genes (Supplementary Fig. 3). Each program utilizes its own criteria of genomic distance, gene pruning algorithm, and biological features. We define disease genes as reported by all four methods and thus derived 405 genes (Supplementary Table 6), of which 109 genes were linked to minimally three AAA trait pairs (Supplementary Fig. 4). Notably, *CELSR2*, *PSRC1*, *LRP1*, and *NOC3L* were each shared among 14 AAA trait pairs or more. Such broad distribution suggests their essential roles in cardiac and metabolic functions. Interestingly, all four genes participate in lipid metabolism; furthermore, all but *NOC3L* have been reported in inflammation ^41, 42^.

Pooling genes from any of the four methods, we discovered their functions were enriched in lipoprotein organization, cholesterol transport, and acylglycerol homeostasis (Supplementary Fig. 5A). Strikingly, cholesterol metabolism was the most enriched pathway across all 21 trait pairs (Supplementary Fig. 5B). When classifying by etiological mechanisms ^18^, the most prominent enrichments appeared in cholesterol metabolism, PPAR pathway in lipid metabolism, TGF-β pathway in inflammation, and ECM-receptor interaction in extracellular matrix dysregulation (Fig. 3C).

Summarizing the shared SNVs and genes, we construct the comorbidity network for AAA, detailing the shared variants and genes for each trait pair (Fig. 4).

**Fig. 4.**
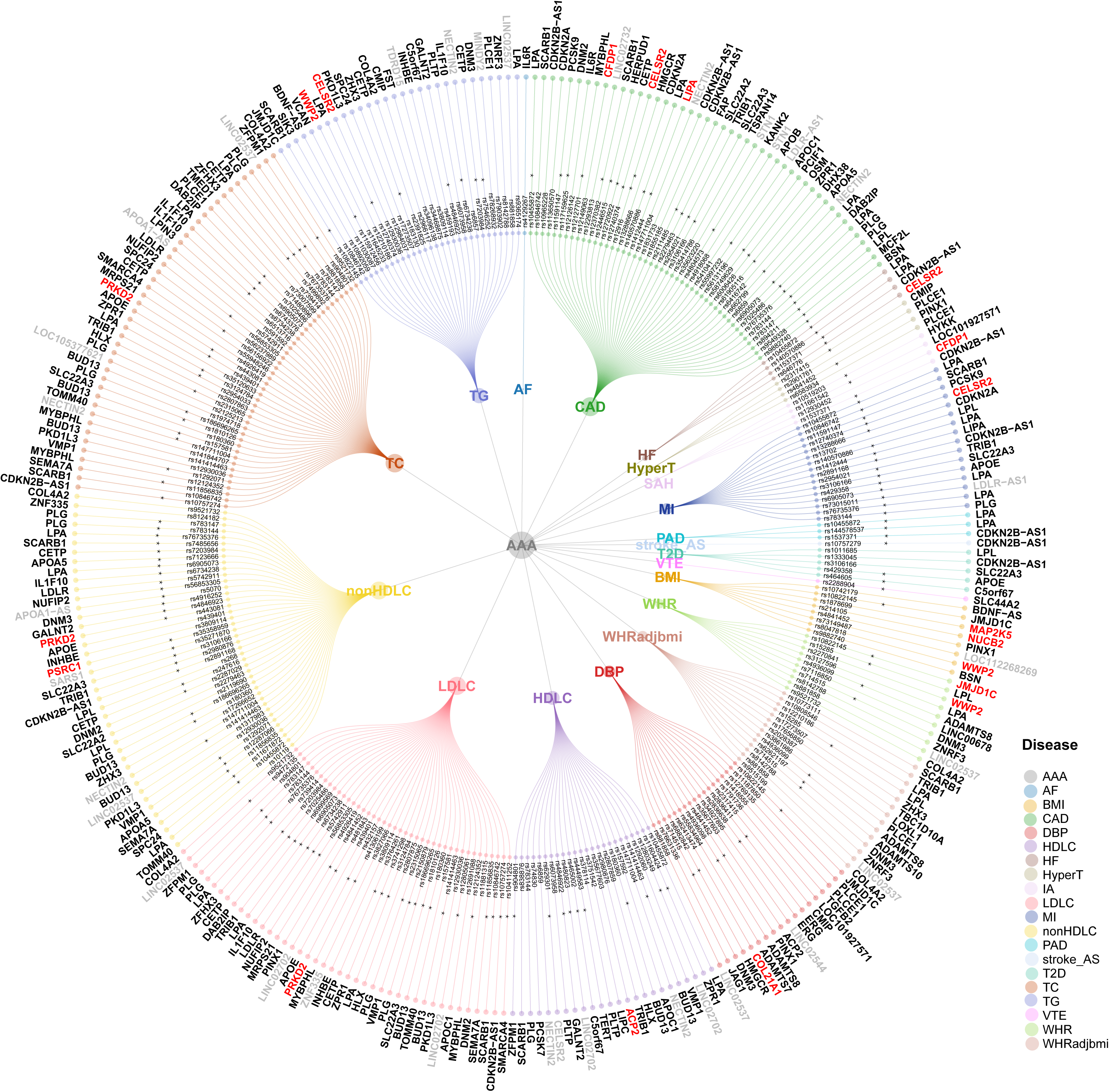
Circular dendrograms presenting shared loci for AAA and CMTs. The inner circle presents independent variants shared between AAA-trait pairs, with 177 shared causal variants marked in asterisks (posterior probability of H4 [PP.H4] > 0.7). The outer circle presents the genes inferred by Annovar for the shared variants. Genes are highlighted by colors to indicate overlap with the four gene identification methods: GCTA, MAGMA, TWAS and SMR, with gray for not identified by any method, black color for identified by at least one method, and red color for identified by all four methods.

### Tissue and cell-type specificity

The shared genes may function in certain tissues and cell types more specifically. We examined it from gene expression in GTEx ^43^ and single-cell transcriptome, as well as heritability in tissue-specific genes and cell type-specific enhancers in CATLAS ^44^. Combing both approaches, we discovered that liver, artery, and adipose tissue (Supplementary Fig. 6), and adipocytes, hepatocytes, fibroblasts, vascular smooth muscle cells, macrophages, and myeloid cells (Supplementary Fig. 7) were significantly enriched across many AAA trait pairs, suggesting them as hubs for cardiac and metabolic functions (Fig. 5). Unique sharing is observed too. As examples, muscle is only enriched by AAA and atrial fibrillation, pituitary and brain are only enriched by AAA and BMI, and pancreas is only enriched by AAA and HDL-C. While fibroblasts are broadly shared across traits, macrophages and hepatocytes are more specific to AAA and lipid traits. Overall, these results align with the genes and pathways, highlighting lipid metabolism and immunity over and again.

**Fig. 5.**
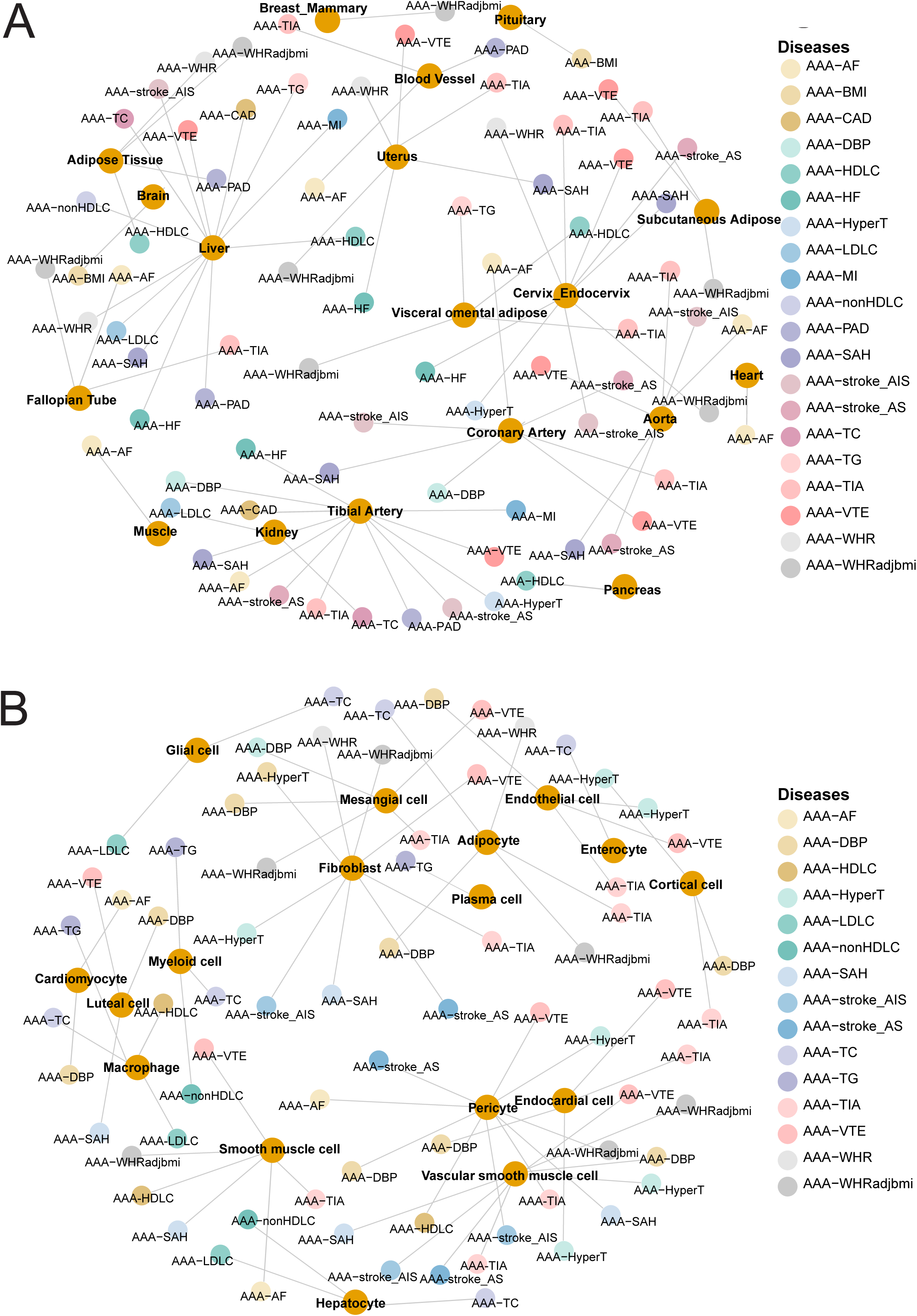
Tissue and cell-type specificity inferred from the shared signals between AAA and CMTs. **A** Enriched tissue types by the heritability or expression of the tissue-specific genes derived from GTEx. **B** Enriched cell types by the heritability of the cell type-specific enhancers derived from CATLAS, or expression of the cell type-specific genes in 11 single-cell transcriptome datasets.

We additionally used SMR ^38^ and TWAS ^37^ to pinpoint gene-tissue effects for each CMT. Collectively, 116 genes were inferred for their directions of effect in tissues (Supplementary Fig. 8 A-B). Here we highlight four most broadly shared genes: *CELSR2*, *PSRC1*, *LRP1*, and *NOC3L.* Both methods detected a negative relationship between CELSR2 expression in the liver with AAA and five other CMTs (Supplementary Fig. 8C). Negative relationships were found for NOC3L expression in the skeletal muscle, and PSRC1 expression in the liver, whole blood, and esophagus mucosa, with AAA and numerous other CMTs. Meanwhile, LRP1 expression in the tibial artery was suggested for a positive relationship with AAA but a negative relationship with CAD.

### Drug for AAA with comorbid conditions

Collectively we identified 405 disease genes shared by AAA and various CMTs. As cardiometabolic disorders often coexist, we used these genes to identify drugs for treating AAA with comorbidities. As such, we utilized a pathway paring score approach developed in our earlier study ^45^ to identify the best matching drugs and disease genes. Briefly, we computed the pathological pathways for each trait pair based on their shared genes, and the pharmacological pathways for each candidate drug based on their affected genes recorded in large drug-gene databases, e.g., DrugCentral ^46^, DGIdb ^47^, and PharmGKB ^48^. The candidate drugs were mainly derived from screening cardiovascular compounds that targeted any of the 405 disease genes. We also supplemented the list with those compounds used in clinical practice or clinical trials for treating AAA. Collectively, 33 candidate drugs distributed in 6 functional classes were examined, namely antihypertension (11 drugs), lipid-lowering (8 drugs), glucose-lowering (3 drugs), antiarrhythmics (1 drug), antithrombosis (4 drugs), and antioxidant (6 drugs) (Supplementary Fig. 9).

The best-matching drugs were defined with pairing scores >= 0.5 (Supplementary Fig. 9). Close to half drugs were suggested for AAA and hypertension, which were distributed in 4 functional categories. Therein amlopidine has the highest pairing score, followed by several antioxidants. Lipid-lowering drugs obtained high pairing scores for various trait pairs. Particularly, simvastatin and lovastatin both achieved high scores for AAA comorbid with CMDs, such as hypertension, MI, subarachnoid hemorrhage, transient ischemic attack, venous thromboembolism, or peripheral artery disease. Interestingly, other lipid-lowering drugs are suggested for AAA with metabolic traits. For example, fenofibrate and gemfibrozil achieved high scores for AAA comorbid with LDL-C, nonHDL-C, triglycerides, or total cholesterol.

Notably, several herb-based antioxidants achieved high scores for various trait pairs too, including resveratrol, a stilbenoid polyphenol naturally enriched in red grapes; tanshinone I, a terpenoid exacted from the dry root of *Salvia miltiorrhiza* (Danshen); and quercetin, a flavonol found in many plants. These herb products have shown potentials in preventing and treating CVDs, including AAA ^49, 50, 51, 52^. Our analysis supports their extended application in treating comorbid conditions in AAA.

## Discussion

In this study, we discovered from GWAS summary statistics extensive genetic associations between AAA and CMTs. Further analyses highlight the pleiotropic variants and genes, the biological pathways and the types of cells and tissues that are shared by the trait pairs. All these findings help to elucidate the common genetic etiology between AAA and cardiometabolic disorders.

We discovered that among all CMDs outside of the aortic aneurysm family (i.e., AAA, TAA, and AA), CAD displays a consistent strong relationship with AAA. For example, it has the second highest genome-wide association (r_g_ = 0.34) and is the only trait with mutual causality with AAA (OR_AAA->CAD_ = 1.10, OR_CAD->AAA_ = 1.23). It shares the largest number of SNVs (N = 46), causal SNVs (N = 30), and disease genes (N = 50) with AAA. The most enriched tissues by these shared signals are artery and liver, consistent with their common features of artery malfunctions and atherosclerosis. Artery-related diseases including peripheral artery disease (r_g_ = 0.33) and subarachnoid hemorrhage (r_g_ = 0.32), and cardiac-function related diseases such as MI (r_g_ = 0.38) and heart failure (r_g_ = 0.30), also displayed top genome-wide associations with AAA, although no causal relationship was found, suggesting other risk factors may have confounded the associations.

Included in this study are the metabolic traits of lipids, adiposity, blood pressures, and glucose. By all levels of our inspection, lipid metabolism is most prominently shared. <u>First</u>, lipid traits are the second strongest causal factor to AAA (OR = 1.46-1.73), next to hypertension, which is a disease rather than a physiological trait. <u>Second</u>, we observed clustering of the shared variants on lipid-related genes, including *LPA*, *CDKN2B-AS1* and others. <u>Third</u>, the most broadly shared genes between AAA and CMTs, i.e., *LRP1*, *PSRC1*, *CELSR2*, and *NOC3L*, are all lipid related. <u>Fourth</u>, cholesterol metabolism appeared as the most significantly enriched biological pathway. <u>Fifth</u>, liver, adipose tissue, hepatocytes, and adipocytes are most broadly and significantly enriched among the AAA-CMT trait pairs. These tissues and cell types are important players in lipid metabolism and regulation. <u>Lastly</u>, lipid-reducing drugs were suggested as strong candidates to treat many AAA with comorbid diseases. These results reinforce the notion that predisposition to lipid malfunction is a strong feature in CMTs ^53^. In comparison, glucose traits demonstrate neither correlation nor causality to AAA. Among the blood pressure traits, only diastolic blood pressure displays a mild correlation (r_g_ = 0.16) and a weak causality (OR_DBP◊AAA_ = 1.05).

Several genes appeared repetitively in our analyses. *LPA* encodes lipoprotein(a), which is pro-atherosclerotic, pro-inflammatory, pro-thrombotic, and anti-fibrinolytic. Substantial evidence suggest that elevated lipoprotein(a) promotes CAD, MI, atherosclerosis, and aortic valve stenosis ^54, 55^. *CDK2B-AS1* encodes a long non-coding RNA that participates in inflammation as well as the metabolism of lipids and carbohydrates, and has been linked to numerous CMDs and immune diseases ^56, 57^. *LRP1* encodes LDL receptor-related protein and plays diverse roles in lipoprotein metabolism, endocytosis, cell growth, cell migration, inflammation, and apoptosis ^42^. Furthermore, CELSR2 and PSRC1, together with SORT1, form a PRSC1-CELSR2-SORT1 axis which has been implicated in various CVDs ^41, 58^. *SORT1* encodes sortilin 1 that functions in lipid metabolism and immune responses, such as V-LDL secretion, LDL-C metabolism, PCSK9 secretion, inflammation, and formation of foam cells ^59^. Finally, NOC3L is involved in adipocyte differentiation and glucose metabolism, and its decreased expression is associated with islet dysfunction ^60^.

We note that various disease genes in lipid metabolism are involved in immune responses too. Indeed, *LPA* is pro-inflammatory ^61^; *CDK2B-AS1* is not only associated with numerous CMDs but also with immune diseases, such as idiopathic pulmonary fibrosis and inflammatory bowel disease ^56, 57, 62^. Interestingly, statins, other than lowering lipids, are found to inhibit inflammation in AAA ^63^.

Indeed, there are abundant immune signals in our results. For example, IL-6 is an important cytokine in CVDs including AAA ^64^. Enhanced IL-6 signaling will over-activate the JAK-STAT pathway, a critical pathway that affects many aspects of the mammalian immune system ^65^. rs6734238 was reported to associate with elevated circulating IL-6 ^66^, whereas our analysis inferred this SNV as causal to AAA, LDL-C, total cholesterol, and triglycerides (Fig. **3**B). We also identified two SNVs in the intronic regions of *IL6R*, rs4129267 and rs12126142, to be shared by AAA with atrial fibrillation and CAD, respectively. Furthermore, our pathway enrichment highlights the TGF-β signaling, which was shared by AAA and 12 CMTs (Fig. **3**C). TGF-β regulates the differentiation and function of leukocytes and controls the type and scope of immune response ^67^. Numerous studies have uncovered its importance in vascular smooth muscle cells (SMCs) and macrophages in the aneurysm development ^68, 69^. SMCs can transdifferentiate to foam cells, a crucial step in atherosclerosis ^70^. In our analysis, both vascular SMCs and macrophages were enriched by several AAA trait pairs. Indeed, various single-cell RNA-sequencing studies suggested them as essential cell types for AAA ^71, 72^. Our analysis reveals this close relationship also in genetic predisposition.

Overall, many of our results recapitulate the relationships of AAA with its risk factors and known disease markers, indicating our results captured the main components of AAA genetics. To confirm this, we analyzed the AAA single trait GWAS loci. Many of the shared lipid-related genes are reproduced, and genes in various pathological mechanisms are connected (Supplementary Fig. 10). The most significantly enriched terms are lipid processes and cholesterol metabolism. The most enriched tissues are liver and blood vessels, and the most enriched cell types are fibroblasts, with a few others showing marginal enrichment, including endothelial cells, stromal cells, mesenchymal stem cells, macrophages, neutrophils, and monocytes. Therefore, the shared signals are the main signals in AAA genetics.

Finally, the shared disease genes are transformed to treatment proposals for treating AAA with comorbid conditions. We believe this drug prioritization strategy can be applied to studying other diseases with comorbidities.

There are several limitations of this study. First, the GWAS data type enables analysis on common SNVs, but omits other variant types such as rare variants, the short insertions and deletions (INDELs), and structural variants (SV). Indeed, our previous whole-genome study identified a list of rare variants with strong predictability to AAA ^16^. Second, CMTs cover a plethora of diseases and physiological traits, and those included in our analysis are only representative. Third, our inference of molecular and cellular mechanisms may be limited by the reference knowledgebases and databases. For example, we only deciphered the directions of effects for a quarter of the disease genes, due to the lack of variant-gene expression models in GTEx. With future improvements in the data types and references, we will gain further power to interpret results and infer genetic mechanisms of AAA and other CMTs.

## Methods

### Study populations

We obtained summary statistics for the multi-ancestry meta-GWAS of AAA (39,221 cases and 1,086,107 controls) from ^18^. Summary statistics for other 32 CMTs were derived from UK Biobank (https://www.ukbiobank.ac.uk/), FinnGen (https://www.finngen.fi/en), and numerous large consortia ^73, 74, 75^. Summary statistics for the counts and percentages of six immune cell traits were derived from the Blood Cell Consortium (BCX) ^76^, including white blood cell (WBC), basophil (BASO), eosinophil (EO), lymphocyte (LYMPH), monocyte (MONO), and neutrophil (NEUT), obtained from 563,085 participants of European ancestry. Information of these GWAS studies is provided in Supplementary Table 1.

### Genome-wide genetic correlation

We computed genome-wide genetic correlation between traits using linkage disequilibrium (LD) score regression (LDSC) ^21^. Briefly, it quantifies the separate contributions of polygenic effects by examining the relationship between LD scores and test statistics of SNVs from GWAS summary results, producing genetic correlation based on the deviation of chi-square statistics from the null hypothesis. LDSC also applies a self-estimated intercept during the analysis to account for shared subjects between studies. The derived estimates range from –1 to 1, with –1 indicating a perfect negative genetic correlation and 1 indicating a perfect positive genetic correlation. We used pre-computed LD scores obtained from ∼1.2 million common SNVs in the well-imputed HapMap3 European ancestry panel. A Bonferroni-corrected *P* value threshold of 0.0015 (0.05/32) was used to define statistical significance.

### Genetic correlation by functional categories

We used LDSC to estimate genetic correlations between traits in 24 functional categories ^77^, e.g., transcribed regions, repressed regions, conserved regions, coding regions, promotors, enhancers, superenhancers, introns, transcription factor binding sites (TFBS), DNaseI digital genomic footprinting (DGF) regions, DNase I hypersensitivity sites (DHSs), fetal DHS, untranslated regions (UTR), and histone marks (H3K4me1, H3K4me3, H3K9ac, and H3K27ac) from the Roadmap Epigenomics Project ^77, 78^. For each functional category, SNVs from the panel of HapMap3 European ancestry were assigned and LD scores were calculated, generating the “baseline model” (https://github.com/bulik/ldsc/wiki/Partitioned-Heritability). We downloaded them as the ldscore reference file to compute heritability enrichment and genetic correlation for the 24 functional categories.

### Mendelian randomization (MR) analysis

We used five MR methods to infer causal relationships between AAA and CMTs: inverse variance weighting (IVW) ^79^, MR-Egger ^80^, weighted median ^81^, MR-APSS ^82^, and CAUSE ^83^.

These methods utilize different assumptions about horizontal pleiotropy. Briefly, IVW assumes mean zero if uncorrelated pleiotropy is present, and such pleiotropy only adds noise to the regression of the meta-analyzed SNV effects with multiplicative random effects ^79^. MR-Egger further allows for the presence of directional (i.e., non-zero mean) uncorrelated pleiotropy and adds an intercept to the IVW regression to exclude such confounding effect ^80^. Weighted median approach provides a robust estimate of causal effects even when up to 50% of genetic variants are invalid ^81^. The recently published MR-APSS accounts for pleiotropy and sample structure, simultaneously ^82^. Specifically, for decomposing the observed SNV effects, a foreground-background model is employed, in which the background model accounts for confounding factors (including correlated pleiotropy and sample structure) hidden in the GWAS summary statistics, and the foreground model performs causal inference while accounting for uncorrelated pleiotropy. CAUSE is a Bayesian MR method accounting for both correlated and uncorrelated pleiotropy ^83^. Compared to the other MR methods, CAUSE further corrects correlated pleiotropy by evaluating the joint distribution of effect sizes from instrumental SNVs, assuming that the ‘true’ causal effect can influence all instrumental SNVs while correlated pleiotropy only influences a subset of them. CAUSE improves the power of MR analysis by including a larger number of LD-pruned SNVs with *P* <= 1 x 10^−3^ and provides a model comparison approach to distinguish causality from horizontal pleiotropy.

For selecting instruments, we used the genome-wide significance threshold *P* = 5 x 10^−8^ for IVW, MR Egger, and Weighted-median, the default threshold *P* = 1 x 10^−3^ for CAUSE, and the default threshold *P* = 5 x 10^−5^ for MR-APSS. We only selected independent SNVs (LD clumping r2 < 0.001 within 1000 Kb using PLINK v1.9 ^84^) based on the European ancestry panel in the 1000 Genomes Project. In each LD block, we chose the variant with the smallest association *P* value with the exposure. Further, we used PhenoScanner (http://www.phenoscanner.medschl.cam.ac.uk/) and GWAS Catalog (https://www.ebi.ac.uk/gwas/) to exclude SNVs associated with the outcome and its risk factors. IVW was used as the primary method and the rest four methods were used as sensitive analysis. A causal estimate was considered significant if it passed the *P* value threshold in the primary analysis, i.e., IVW, and displayed consistent direction of effect in all five MR methods. We used Cochran’s Q-test to check for heterogeneity and MR-Egger intercept for horizontal pleiotropy. These MR analyses were performed in the R packages TwoSampleMR^85^, MRAPSS ^82^ and CAUSE ^83^.

### Cross-trait meta-analysis

To identify pleiotropic loci shared between two traits, we performed cross-trait meta-analysis of GWAS summary statistics using MTAG ^22^. MTAG applies generalized inverse-variance-weighted meta-analysis for multiple traits; in addition, it accommodates potential sample overlap between GWAS. Its key assumption is that all SNVs share the same variance-covariance matrix of effect sizes among traits. As initially described ^22^, MTAG is a consistent estimator whose effect estimates have a lower genome-wide mean squared error than the corresponding single-trait GWAS estimates. In addition, association statistics from MTAG also yield stronger statistical power and little inflation of the FDR for each analyzed trait with high correlation ^22^.

As the assumptions in MTAG, i.e., equal SNV heritability for each trait and the same genetic covariance between traits, could be violated, we performed cross-phenotype association analysis (CPASSOC) ^23^ across traits as a sensitivity analysis. CPASSOC integrates GWAS summary statistics from multiple traits to detect shared variants while controlling population structure and cryptic relatedness ^23^. It provides two test statistics, SHom and SHet. SHom is based on the fixed-effect meta-analysis and can be viewed as the maximum of weighted sum of trait-specific genetic effects. It is less powerful under the presence of between-study heterogeneity, which is common when meta-analyzing multiple traits. SHet is an extension to SHom with improved power that allows for heterogeneous effects of a trait from different study designs, environmental factors, or populations, as well as heterogeneous effects for different phenotypes, which is more common in practice. SHet was thus adopted for our analysis. We applied PLINK clumping to obtain the independent SNVs (parameters: --clump-p1 5 x 10^−8^ --clump-p2 1 x 10^−5^ ---clump-r2 0.1 --clump-kb 1000). Significant pleiotropic SNVs were defined as variants with *P* values in both GWAS studies and *P* value in the meta-analysis (i.e., *P_MTAG_* & *P_CPASSOC_*) < 5 x 10^−8^. We used ANNOVAR for functional annotation of the variants identified by MTAG and CPASSOC. The shared SNVs are visualised in a circular dendrogram using the R package ggraph.

### Fine-mapping credible set analysis

We identified a 99% credible set of causal variants by FM-summary (https://github.com/hailianghuang/FM-summary) ^24^, a Bayesian fine-mapping method. For each shared SNV identified in the cross-trait meta-analysis, we extracted variants within 500K bp around the index SNV as input for FM-summary. FM-summary set a flat prior and produced a posterior inclusion probability (PIP) of a true association between a phenotype and a variant using the steepest descent approximation. A 99% credible set is equivalent to ranking the SNVs from largest to smallest PIPs and taking the cumulative sum of PIPs until it is at least 99%.

### Colocalization analysis

We used the R package coloc ^25^ to determine whether the association signals for AAA and CMTs co-localize. For each of the 203 shared SNVs between traits, we extracted the variants within 500 Kb of the index SNV and calculated the probability that the two traits share one common causal variant (H4). Loci with a probability greater than 0.7 were considered to colocalize. We estimated the posterior probability (PP) of multiple traits sharing the same SNV using a Bayesian divisive clustering algorithm implemented by HyPrColoc ^26^ (v.1.0.0) in R v.4.2.3.

### Gene-based association analysis

We used TWAS ^37^, SMR ^38^, MAGMA ^39^, and GCTA-fastBAT ^40^ to identify genes shared by AAA trait pairs. Input files for all four gene-level analyses were the complete GWAS summary statistics from MTAG in the meta-analysis. In each method, the *P* value threshold was adjusted by Bonferroni correction.

TWAS identifies tissue-specific gene-trait associations by integrating GWAS with *cis*-SNVs based gene expression model ^37, 86^. We conducted TWAS using the FUSION software ^37^ based on 43 post-mortal tissue expression profiles in GTEx (version 6) ^43^.

Summary-data-based Mendelian Randomization (SMR) analysis integrates GWAS and eQTL studies to identify genes whose expression levels are associated with a complex trait due to pleiotropy or causality ^38^. A significant SMR association could be explained by a causal effect (i.e., the causal variant influences disease risk via changes in gene expression), pleiotropy (i.e., the causal variant has pleiotropic effects on gene expression and disease risk), or linkage (i.e., different causal variants exist for gene expression and disease). SMR implements the HEIDI-outlier test to distinguish pleiotropy from linkage. We implemented SMR using *cis*-eQTL summary data for whole blood from eQTLGen ^87^, a meta-analysis of 31,684 blood samples, and from GTEx V8 for 9 relevant tissues, including artery aorta, adipose subcutaneous, artery coronary, artery tibial, heart atrial appendage, heart left ventricle, kidney cortex, liver, and whole blood. Genes associated with AAA trait pairs were defined as *P*_SMR_ passing the Bonferroni-corrected thresholds and *P*_HEIDI_ >0.05.

MAGMA^39^ (Multi-marker Analysis of GenoMic Annotation) is a fast and flexible method that uses a multiple regression approach to properly incorporate LD between markers and detect multi-marker effects. We ran MAGMA with default parameters, with the European ancestry panel in the 1000 Genomes Project (Phase 3) as the LD reference.

We applied a fourth approach, GCTA-fastBAT ^40^, a fast set-based association analysis. In brief, it calculates the association *P* value for a set of SNVs from an approximated distribution of the sum of χ2-statistics over all SNVs using GWAS summary data and LD correlations from a reference sample set with individual-level genotypes ^40^. We used the European ancestry panel in the 1000 Genomes Project (Phase 3) as the LD reference.

### Stratified LD score regression for tissue and cell type specificity

We used LD score regression applied to specifically expressed genes (LDSC-SEG) ^88^ for tissues or cell types to test for heritability enrichment. For tissues, pre-computed LD scores from GTEx ^89^, which contained gene expression data for 53 tissues, were provided by LDSC-SEG and used in our analysis. We also obtained the activity profile of candidate cis-regulatory elements (cCREs) in 222 cell types from CATLAS ^44^. We mapped the genotypes of European ancestry in the 1000 Genomes Project to the cell type-specific cCREs and calculated the cell type-specific ldscore. We applied FDR correction for each dataset respectively to account for multiple testing, and considered FDR corrected *P* < 0.05 as significant.

### GTEx Tissue Specific Expression Analysis (TSEA)

We conducted tissue-specific expression analysis (TSEA) ^90^ on the shared genes against the RNA-Seq data in GTEx, which contained gene expression profiles of 1,839 samples from 45 different tissues derived from 189 post-mortem subjects. We merged the shared genes for each trait pair from the four gene-based analysis to derive a collection of shared genes. Hypergeometric tests are used to determine if tissue-specific genes are enriched in the input genes. We used Benjamini–Hochberg correction to account for multiple testing (FDR < 0.05).

### Cell-type-specific enrichment analysis (CSEA)

We performed cell-type specific enrichment (CSEA) on the shared genes using WebCSEA ^91^. WebCSEA provides a gene set query against tissue-cell-type (TCs) expression signatures of 11 single-cell gene expression datasets ^92, 93, 94, 95, 96, 97, 98^. Specifically, Dai et al ^91^ collected more than 5.5 million cells from 111 tissues and 1,355 TCs, filtered out the low expression genes, and used an in-house t-statistic-based method “deTS” to train the tissue-cell-type signature genes. Genes with the top 5% t-statistic scores in focal cell type are defined as the cell-type-specific genes. We conducted Fisher’s exact test to assess whether the shared genes for each trait pair is overrepresented with the cell type-specific genes.

### Over-representation enrichment analysis

We used the “clusterProfiler” package ^99^ to perform GO (Gene Ontology) and KEGG (Kyoto Encyclopedia of Genes and Genomes) pathway enrichment analysis on the genes. Benjamini – Hochberg procedure was used to account for multiple testing (FDR < 0.05).

### Drug target analysis

We defined the genes identified by all four gene-based analyses as the disease genes for each AAA trait pair and thus obtained 405 genes collectively. For deriving the drugs that match best with the genes, we leveraged the biological pathway analysis. First, we applied clusterProfiler to the shared genes to compute the pathological pathways enriched for each AAA trait pair. Next, we carried multiple steps to derive candidate drugs for scrutinization: (1) queried three large drug-gene databases, DrugCentral ^46^, DGIdb ^47^, and PharmGKB ^48^, for drugs that target any of the 405 candidate genes. This initial screening led to about 1,200 compounds, most of which were initially designed for treating cancer; (2) limited the compounds to those already in use by standard clinical practices for treating cardiovascular diseases, which drastically shrank the list to 21 drugs; and (3) supplemented the list with 12 drugs used in clinics or proposed by clinical trials for treating AAA, such as amlodipoine, pitavastatin, and vitamin E. Collectively, 33 candidate drugs were derived. Then, for each drug, we queried the three drug-gene databases again for all their affected genes and computed their enriched pharmacological pathways in clusterProfiler. Finally, we calculated the pairing scores between the pathological pathways of the cross-trait and the pharmacological pathways of the drug ^45^.

## Code availability

LDSC: https://github.com/bulik/ldsc;

PLINK:https://www.cog-genomics.org/plink/1.9;

LAVA: https://github.com/josefin-werme/lava;

TwoSampleMR:https://mrcieu.github.io/TwoSampleMR/;

CAUSE: https://jean997.github.io/cause;

MR-APSS: https://github.com/YangLabHKUST/MR-APSS;

MR-BMA: https://github.com/verena-zuber/demo_AMD;

MTAG: https://github.com/JonJala/mtag;

CPASSOC: http://hal.case.edu/~xxz10/zhu-web/;

Coloc: https://github.com/chr1swallace/coloc;

HyPrColoc: https://github.com/cnfoley/hyprcoloc;

FM-summary: https://github.com/hailianghuang/FM-summary;

FUSION: http://gusevlab.org/projects/fusion/;

MAGMA: https://ctg.cncr.nl/software/magma;

SMR: https://cnsgenomics.com/software/smr/#Overview;

GCTA-fastBAT: https://yanglab.westlake.edu.cn/software/gcta/#fastBAT;

TSEA: http://doughertylab.wustl.edu/tsea/;

FUMA: https://fuma.ctglab.nl/;

WebCSEA: https://bioinfo.uth.edu/webcsea/

## Data Availability

All data produced in the present study are available upon reasonable request to the authors

## Acknowledgements

This study was supported by the National Natural Science Foundation of China (No. 32270626), Greater Bay Area Research Institute of Precision Medicine (Guangzhou) Research Grants (I0005, R2001). We thank members of the Laboratory of Intelligent Computing in Biomedicine in the Greater Bay Area Institute of Precision Medicine (Guangzhou) for insightful discussions and suggestions.

## Author Contributions

CP, SZ and PST collectively designed the study. SZ and CP performed bioinformatic and statistical analyses and generated the figures and tables. PST and CP provided critical biological insight in result interpretation. CP and SZ drafted the manuscript. All authors critically reviewed the manuscript.

## Competing interests

All authors declare no competing interests.

---

**Supplementary Figure 1.**
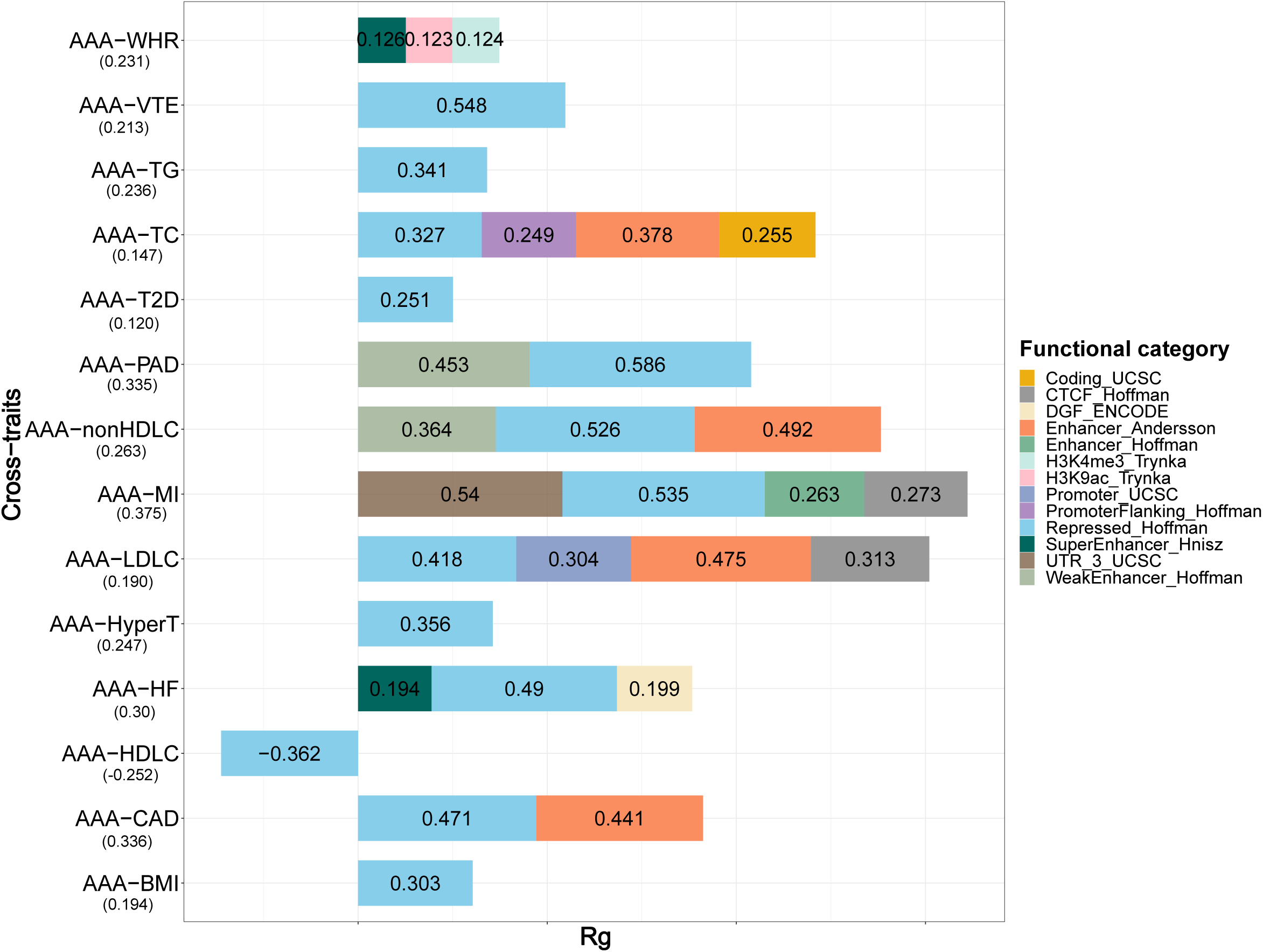
Partitioned genetic correlation between AAA and CMTs. 24 types of genomic functional elements were interrogated, as labeled on the left. The genome-wide correlation was depicted in parenthesis under each trait/disease pair. The bar plot shows functional categories with different correlations than the genome-wide correlation, as defined by a difference r_g_ ≥ |0.1| to the genome-wide correlation value. Colors represent different functional categories.

**Supplementary Figure 2.**
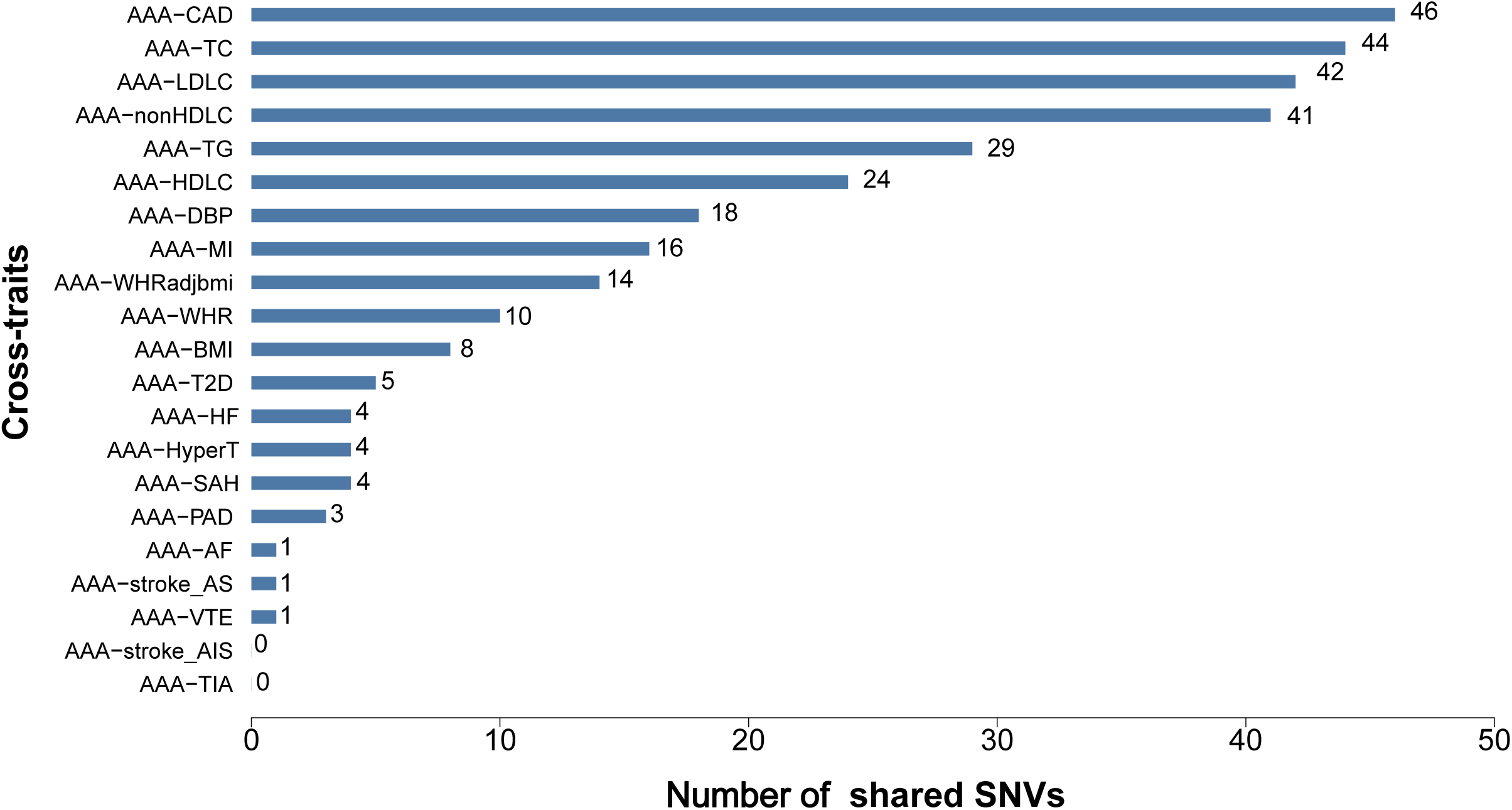
Number of shared SNVs between AAA and CMTs via MTAG and CPASSOC.

**Supplementary Figure 3.**
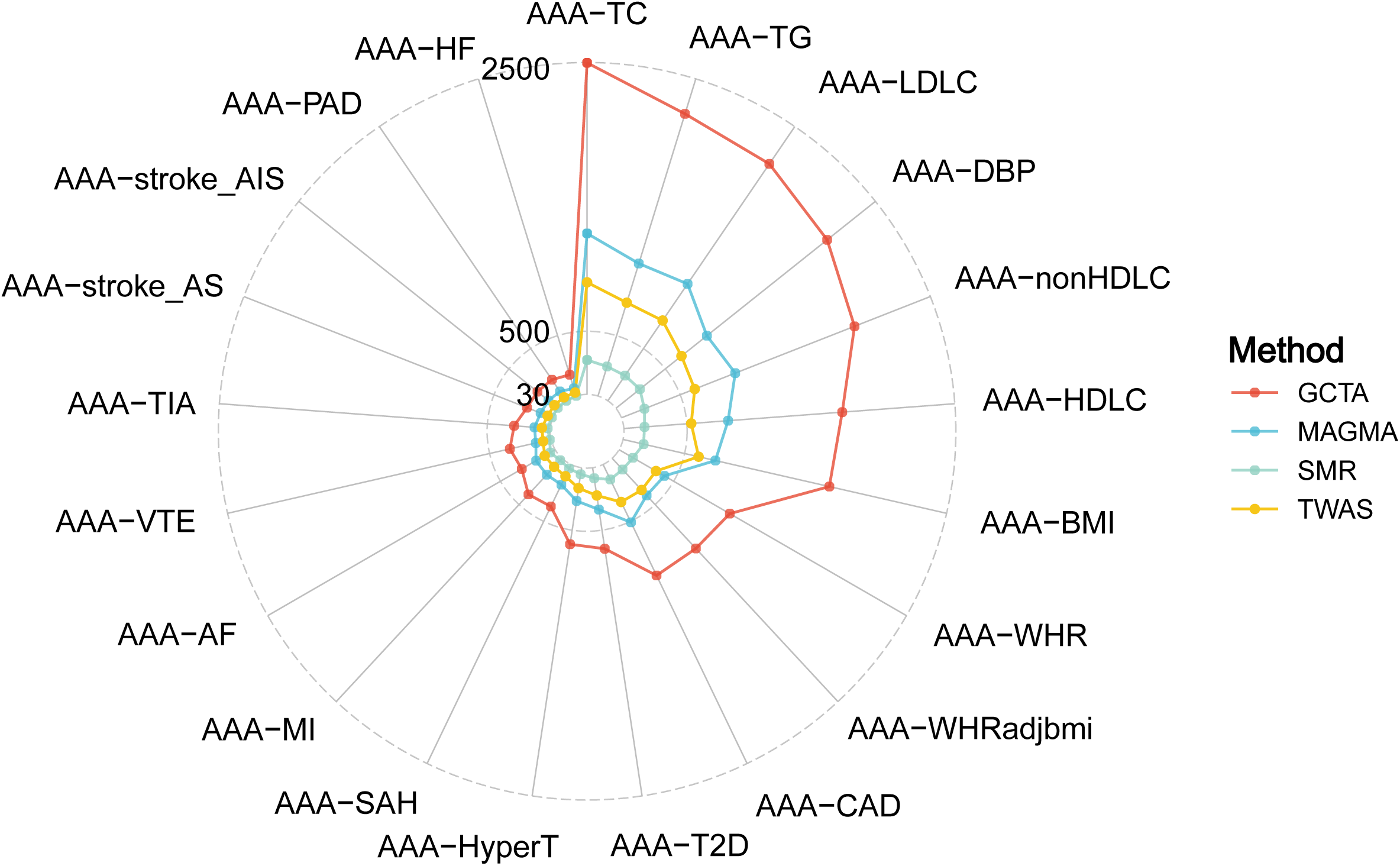
Number of genes for each trait pair identified by four methods: GCTA, MAGMA, TWAS and SMR. Each method is represented by one color. The numbers of identified genes are marked on each tile.

**Supplementary Figure 4.**
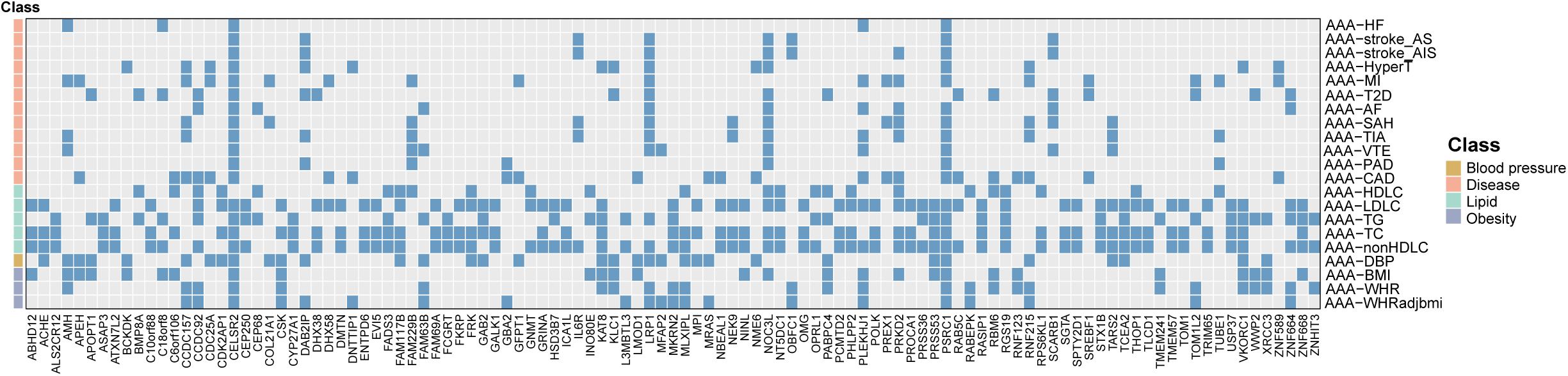
Genes identified by all four gene-based analysis methods and shared by minimally three AAA trait pairs. The four gene analysis methods are: GCTA, MAGMA, TWAS, and SMR. Genes are labeled on the bottom, and the trait pairs are labeled on the right. Blue color indicates presence of the gene.

**Supplementary Figure 5.**
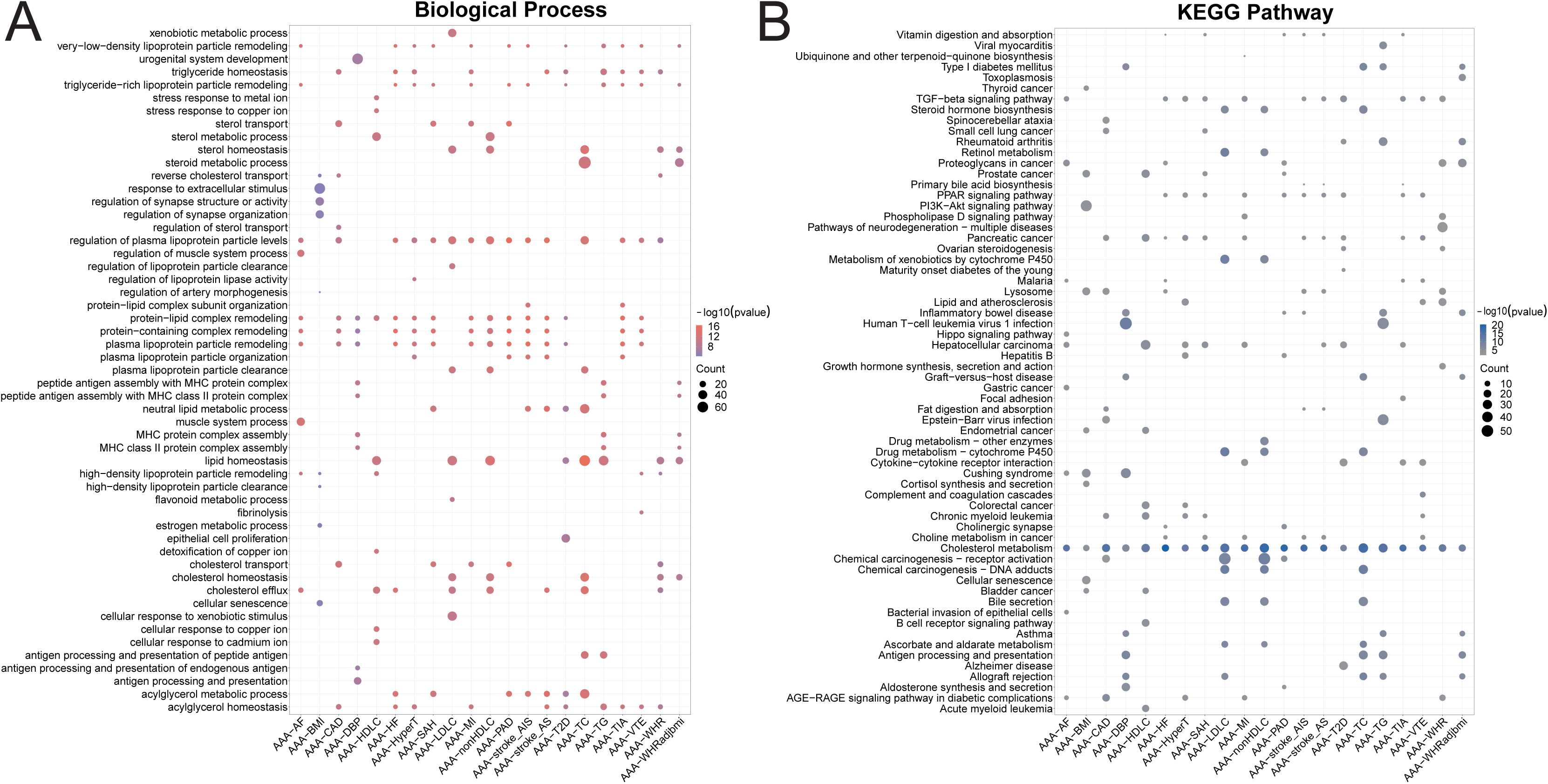
A GO term (biological process) enrichment for AAA and CMTs. B KEGG pathway enrichment for AAA and CMTs. Genes were derived from the union of four gene analysis methods: GCTA, MAGMA, TWAS and SMR. The top 10 enriched pathways passing *P*_adj_ < 0.05 in each trait pair were included.

**Supplementary Figure 6.**
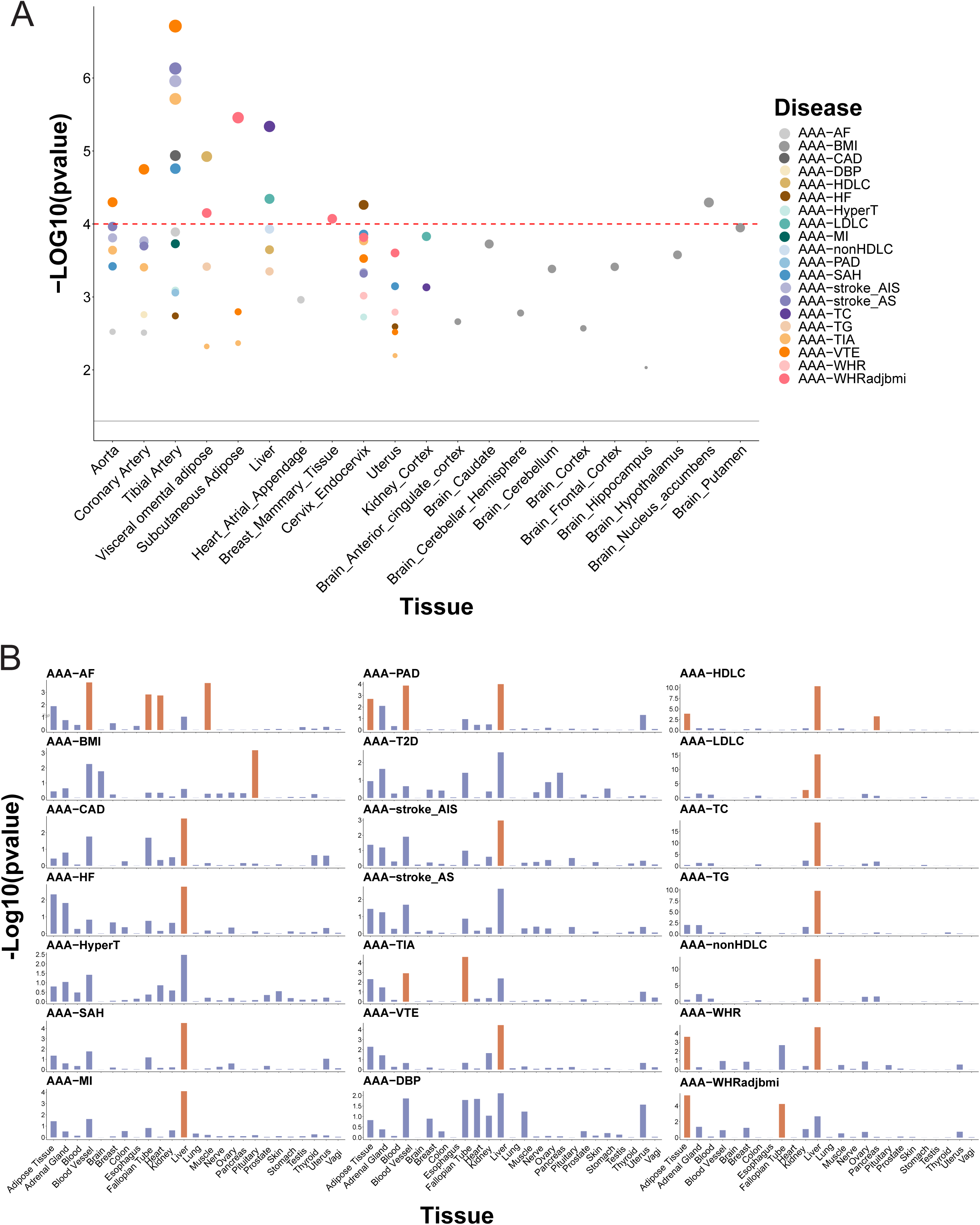
Tissue type enrichment of the shared signals between AAA and CMTs. **A** Enriched by heritability of the tissue-specific genes derived from GTEx. **B** Enriched by the tissue-specific expression in GTEx. Brown represents significant enrichment after Bonferroni correction. Blue represents enrichment result not passing the Bonferroni correction.

**Supplementary Figure 7.**
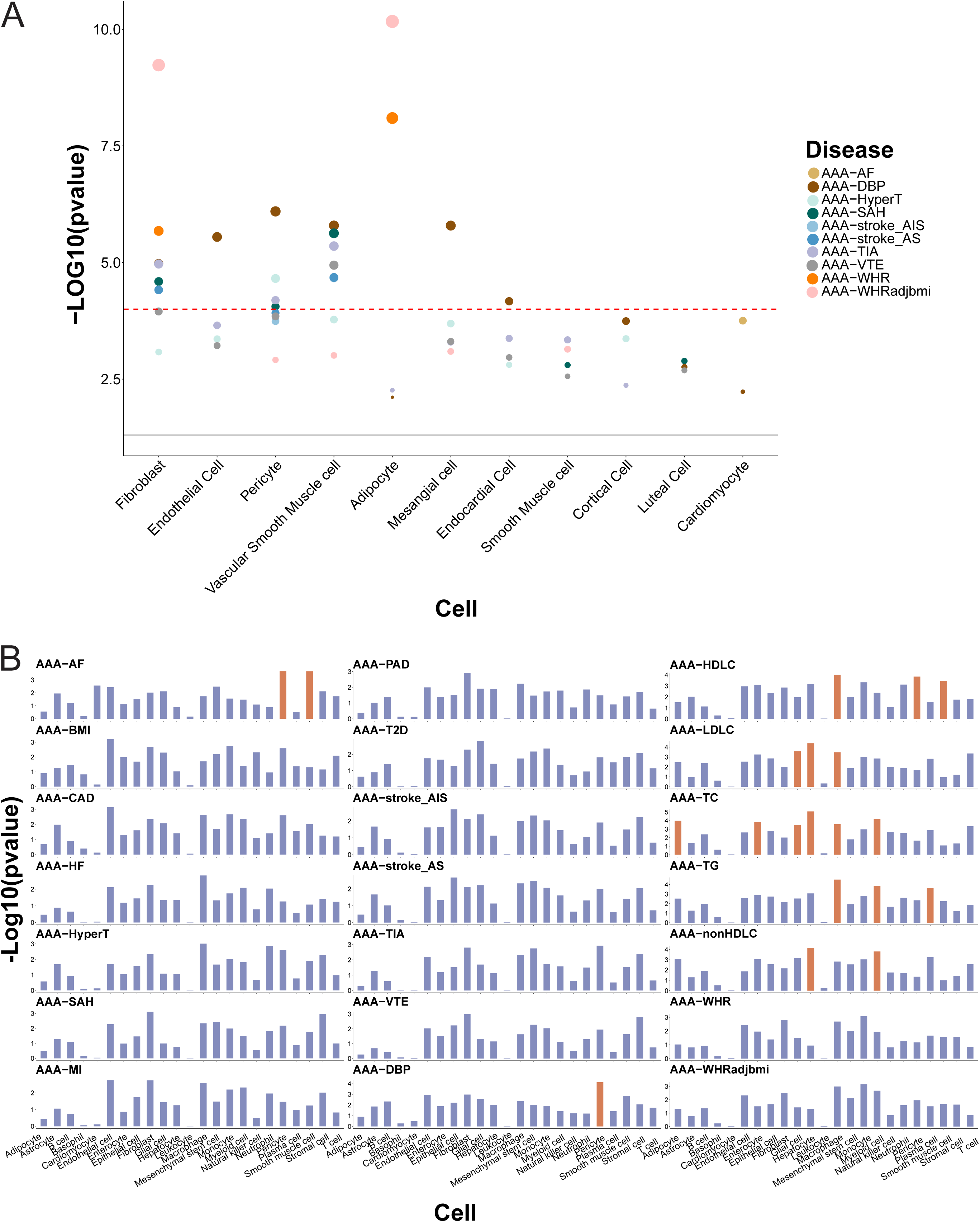
Cell type enrichment of the shared signals between AAA and CMTs. **A** Enriched by heritability of the cell type-specific enhancers derived from CATLAS. **B** Enrichment by cell type-specific expression by referencing to 11 single-cell transcriptome datasets. Brown represents significant enrichment after Bonferroni correction. Blue represents enrichment result not passing the Bonferroni correction.

**Supplementary Figure 8.**
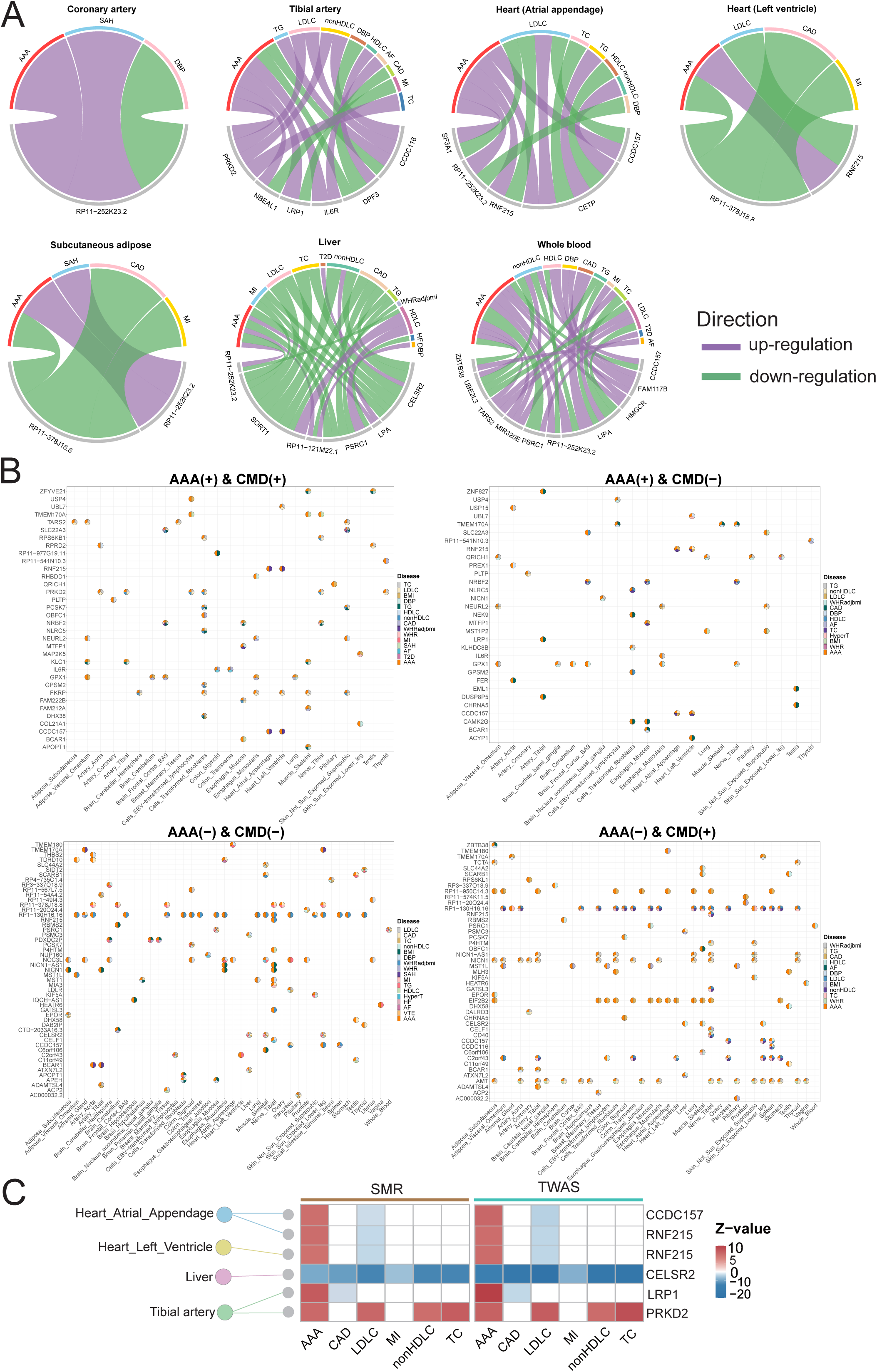
Direction of effect by the genes on the CMTs in the tissue context. **A** SMR-derived directions of effect, inferred by the top causal eQTLs. Traits are marked in the upper half, and the genes are marked in the lower half. Lines within the circles connects genes and the traits. The line color indicates positive (purple) or negative (green) correlation, and the width indicates strength of association derived from Z-scores. Nine tissues impacted the most by the shared signals between AAA and CMTs were examined. **B** Fusion-derived directions of effect, utilizing the model of cis-SNVs on gene expression. 49 tissues from GTEx were interrogated. **C** Overlapped results with *cross trait – tissue - gene* by the two methods are presented.

**Supplementary Figure 9.**
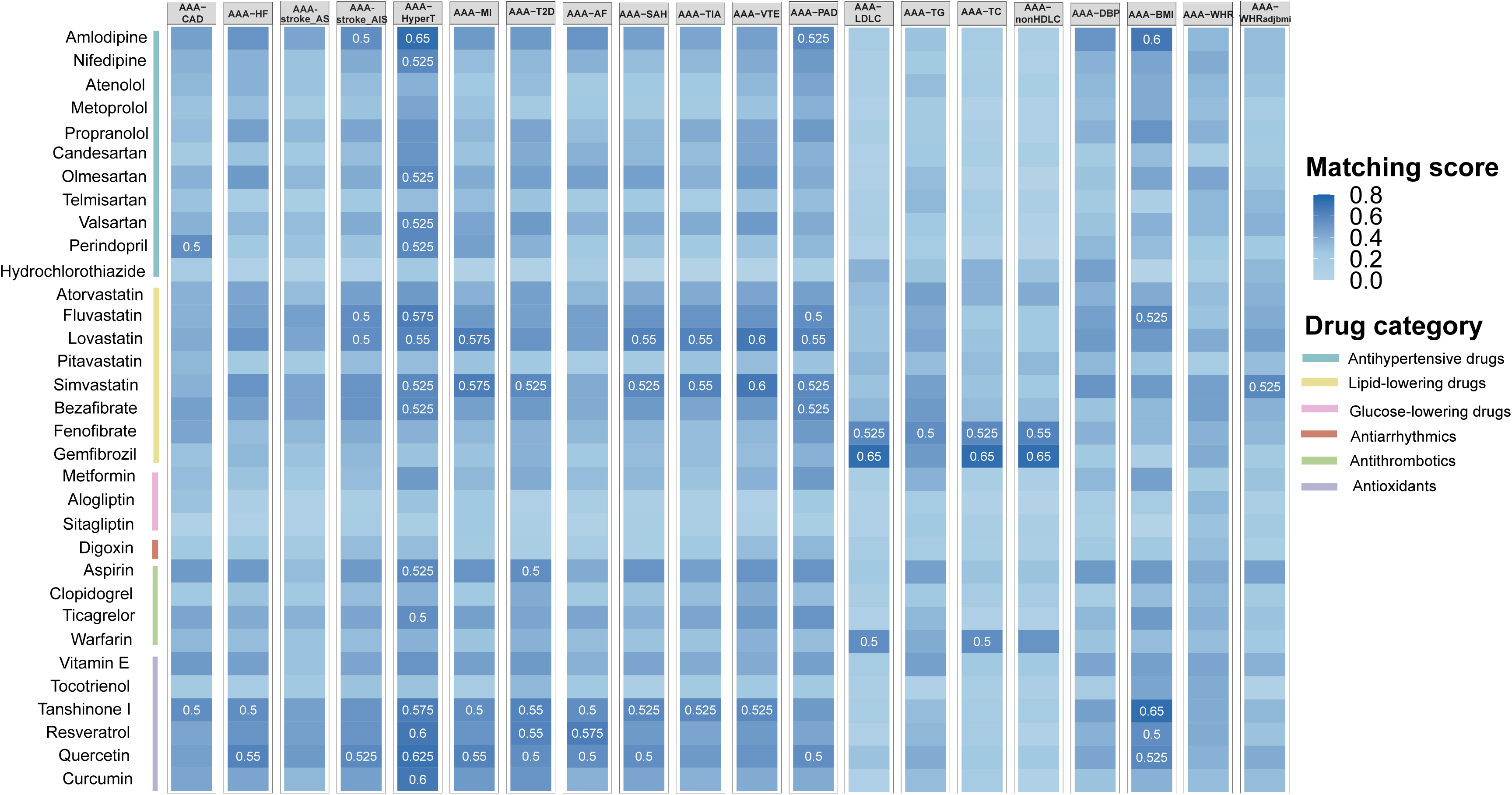
Pairing scores for evaluating matching between disease pathology and drug pharmacology. Matching scores greater than 0.5 and the top ranked in each cross-trait are labeled.

**Supplementary Figure 10.**
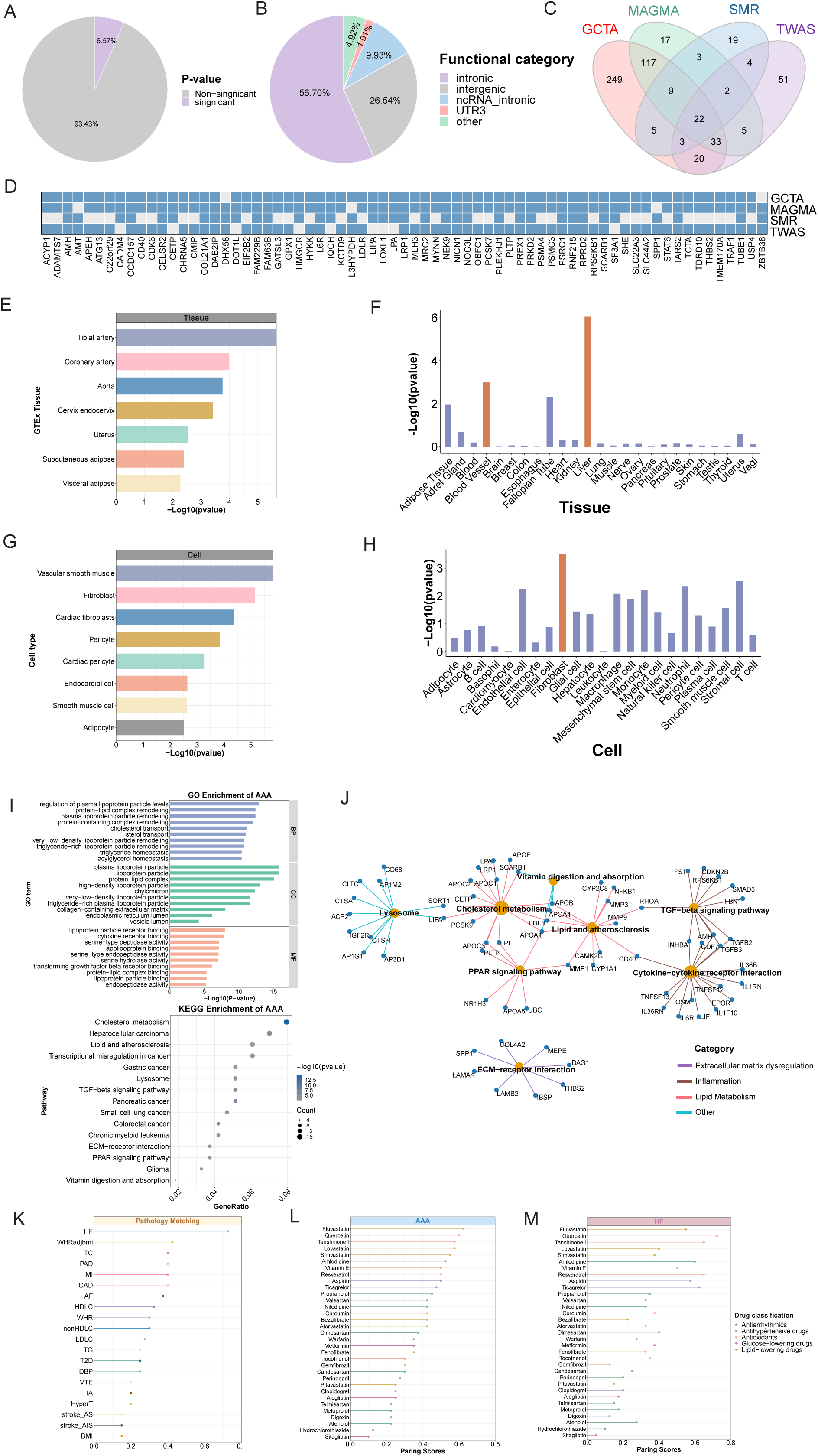
Interpretation of AAA-associated variants identified by GWAS. **A** Distribution of GWAS *P* values. **B** Genic locations of the significant SNVs (GWAS *P* < 5 × 10^-8^). **C** AAA-related genes identified by four gene analysis methods. **D** AAA-related genes identified by minimally three of the four gene analysis methods. **E** Tissue enrichment based on SNV heritability in tissue-specific genes defined in GTEx, computed by S-LDSC. **F** Tissue enrichment based on tissue-specific genes defined in GTEx, computed by Tissue Specific Expression Analysis (TSEA). Orange represents significantly enriched tissues passing the Bonferroni corrected *P* value threshold. **G** Cell type enrichment based on SNV heritability in cell type-specific enhancers from CATLAS, using S-LDSC. **H** Cell type enrichment based on cell type-specific genes defined in various single-cell transcriptome studies, computed by Cell Specific Expression Analysis (CSEA). **I** GO and KEGG enrichment analyses based on the genes identified by any of the four gene analysis methods. **J** Interactions among the genes (blue dots) from the 8 enriched pathways (orange dots) in KEGG enrichment analysis. The colors of the lines represent different functional categories. **K** Pairing scores for the matching between pathological pathways of AAA and CMTs. **(L)** Pairing scores for the matching between AAA pathological pathways and drug pharmacological pathways. **M** Pairing scores for the matching between heart failure and drug pharmacological pathways.

## Notes

### Competing Interest Statement

The authors have declared no competing interest.

### Author Declarations

CVDKP,MEGASTROKE, UK Biobank, CARDIoGRAMplusC4D, Biobank Japan, DIAMANTE, CKDGen, AFGen Consortium, CDKP, FinnGen, GWAS Catalog, MAGIC, GLGC, ICBP, GIANT, Blood Cell Consortium (BCX)

